# Built Environment Features Obtained from Google Street View Are Associated with Coronary Artery Disease Prevalence: A Deep-Learning Framework

**DOI:** 10.1101/2023.03.28.23287888

**Authors:** Zhuo Chen, Yassin Khalifa, Jean-Eudes Dazard, Issam Motairek, Sanjay Rajagopalan, Sadeer Al-Kindi

**Author notes:** **<u>Corresponding Authors</u>** Sadeer Al-Kindi, MD, FACC, Assistant Professor of Medicine, University Hospitals Harrington Heart and Vascular Institute, Case Western Reserve University School of Medicine, 11100 Euclid Ave, Cleveland, OH 44106, OR Sanjay Rajagopalan, MD, FACC, Herman K Hellerstein Professor of Cardiovascular Research, Director, Cardiovascular Research Institute, University Hospitals Harrington Heart and Vascular Institute, Case Western Reserve University School of Medicine, 11100 Euclid Ave, Cleveland, OH 44106. Contributed Equally (Sanjay Rajagopalan, Sadeer Al-Kindi). **<u>Funding</u>:** This work was funded by the National Institute on Minority Health and Health Disparities Award # P50MD017351 and 1R35ES031702-01 awarded to Dr. Rajagopalan.

## Abstract

**Background:** Built environment plays an important role in development of cardiovascular disease. Tools to evaluate the built environment using machine vision and informatic approaches has been limited. We sought to investigate the association between machine vision-based built environment and prevalence of cardiometabolic disease in urban cities.

**Methods:** This cross-sectional study used features extracted from Google Street view (GSV) images to measure the built environment and link them with prevalence of cardiometabolic disease. Convolutional neural networks, light gradient boosting machines and activation maps were utilized to predict health outcomes and identify feature associations with coronary heart disease (CHD). The study obtained 0.53 million GSV images covering 789 census tracts in 7 cities (Cleveland, OH; Fremont, CA; Kansas City, MO; Detroit, MI; Bellevue, WA; Brownsville, TX; and Denver, CO). Analyses were conducted from February 2022 to December 2022. We used census tract-level data from the Centers for Disease Control and Prevention’s PLACES dataset. Main outcomes included census tract-level estimated prevalence of CHD based on GSV built environment features.

**Results:** Built environment features extracted from GSV using deep learning predicted 63% of the census tract variation in CHD prevalence. The ExtraTrees Regressor achieved the best result among all models with the lowest average mean absolute error of 1.11% and Root mean square of error of 1.58. The addition of GSV features outperformed and improved a model that only included census-tract level age, sex, race, income and education. Activation maps from the features revealed a set of neighborhood features represented by buildings and roads associated with CHD prevalence.

**Conclusions:** In this cross-sectional study, a significant portion of CHD prevalence were explained by GSV-based built environment factors analyzed using deep learning, independent of census tract demographics. Machine vision enabled assessment of the built environment could help play a significant role in designing and improving heart-heathy cities.

## Introduction

Coronary heart disease (CHD) accounts for over 50% of mortality from heart disease in the United States, responsible for nearly 400,000 deaths in 2020^1^. Despite advances in prevention and treatment over the past decade in the United States^2^ CHD remains the leading cause of death in the United States since 1950 with increasing evidence for non-conventional risk factors playing a large than anticipated role than previously suspected^1,3^.

Socioenvironmental factors are amongst the leading non-traditional risk factors increasingly implicated in CHD development^4–6^. These factors include social determinants such as race, income, education, and culture as well as the factors in the built environment and factors in the ambient environment such as noise, and air pollution all of which have been to exert significant effects on CHD. ^5–8^

Large-scale integrated assessment of the environment at the neighborhood can facilitate rapid and complete assessment of its impact on CHD. Such data is however scarce, partly because of the costly and time-consuming nature of neighborhood audits, and inconsistent measurements and standards for data collection. Machine vision approaches such as Google Street View (GSV) has become an increasingly popular approach for virtual neighborhood audits since its launch in 2007. GSV image coverage has been consistently expanding in recent years achieving almost full coverage in the United States^9^. Previous studies have shown GSV results are comparable to field assessments and have been used to assess the built environment features such as greenspace^10,11^, buildings^12^, and roads^13^.

GSV images further become a favored data source for large-scale studies due to the open availability of such data, arguably the largest compendium of machine vision enabled assessment of large tracts of the earth, and the standardized approaches used. Deep learning approaches such as convolutional neural networks (CNN) have been widely used in many studies and applications, given their excellent performance in tasks such as image classification, object detection, and image segmentation^14^. The use of such approaches to rapidly assess and extract built environment features from GSV images using deep learning can help facilitate integrated assessment and capture other aspects that may not be otherwise included. The goal of this study is to use GSV images to assess built environment and use them to estimate CHD prevalence at the census tract level.

## Methods

### Data source for coronary heart disease

The prevalence of census-tract coronary heart disease (CHD) was obtained from the CDC PLACES, a project that provided chronic disease risk factors, health outcomes, and clinical preventive services. This project, is a collaboration between the Centers for Disease Control and Prevention (CDC), the Robert Wood Johnson Foundation, and the CDC Foundation, measures CHD prevalence using data from Behavioral Risk Factor Surveillance System (BRFSS), where people aged ≥18 are surveyed to report whether or not they have been told by a doctor, nurse, or other health professional that they had angina or coronary heart disease. We collected the CHD prevalence data for 789 census tracts in 7 cities: Bellevue, WA; Brownsville, TX; Cleveland, OH; Denver, CO; Detroit, MI; Fremont, CA; and Kansas City, KS.

### Google street view data

Environment information was derived from approximately 0.53 million GSV images for the 7 cities (143K for Detroit, 59K for Kansas City, 70K for Cleveland, 65K for Brownsville, 38K for Fremont, 35K for Bellevue, and 120K for Denver). The GSV images were downloaded via Google Street View Static Application Programming Interface (API) from 2020-2021. GSV API provides users with street-level panoramic imagery which captures the visual domain of pedestrians in thousands of cities worldwide. The GSV images of each census tract were downloaded in a grid pattern in the corresponding tract with an interval of 100m. At each location where GSV images were retrieved, four images were gathered from different directions (i.e., the cardinal directions: N, E, S, and W.), which composes a panoramic view of the surroundings at that location. When latitude and longitude coordinates are provided, the API searches within a 50-meter radius for a photograph closest to this location. The API would not return any images if no available images could be found.

To process these images and gain environment information from them, a pre-trained deep convolutional neural network (DCNN) Place365 CNN ^15^ was used as the feature extractor to obtain the deep features of the image. Here, the deep features are the outputs of the deep layers in the hierarchy of the network. Compared with the shallow features in the shallow layers, these deep features represent the semantic information of the GSV images. Details of how the extraction was performed can be found in e Figure 1 in the Supplement. We used Place365 CNN as the feature extractor because the images trained on Place365 CNN are more similar to that of GSV. Place365 CNN was trained on the subset of Places Database, which contains more than 10 million images consisting of 400+ unique scene categories such as towers, soccer fields, streets, swimming pools, and train station platforms. Compared with the ImageNet database, the diversity of environmental features found in the Places Database was believed to be representative of what is contained in GSV images. Through feature extraction, we obtained 4096 features representing the average built environment information for each census tract.

**Figure 1.**
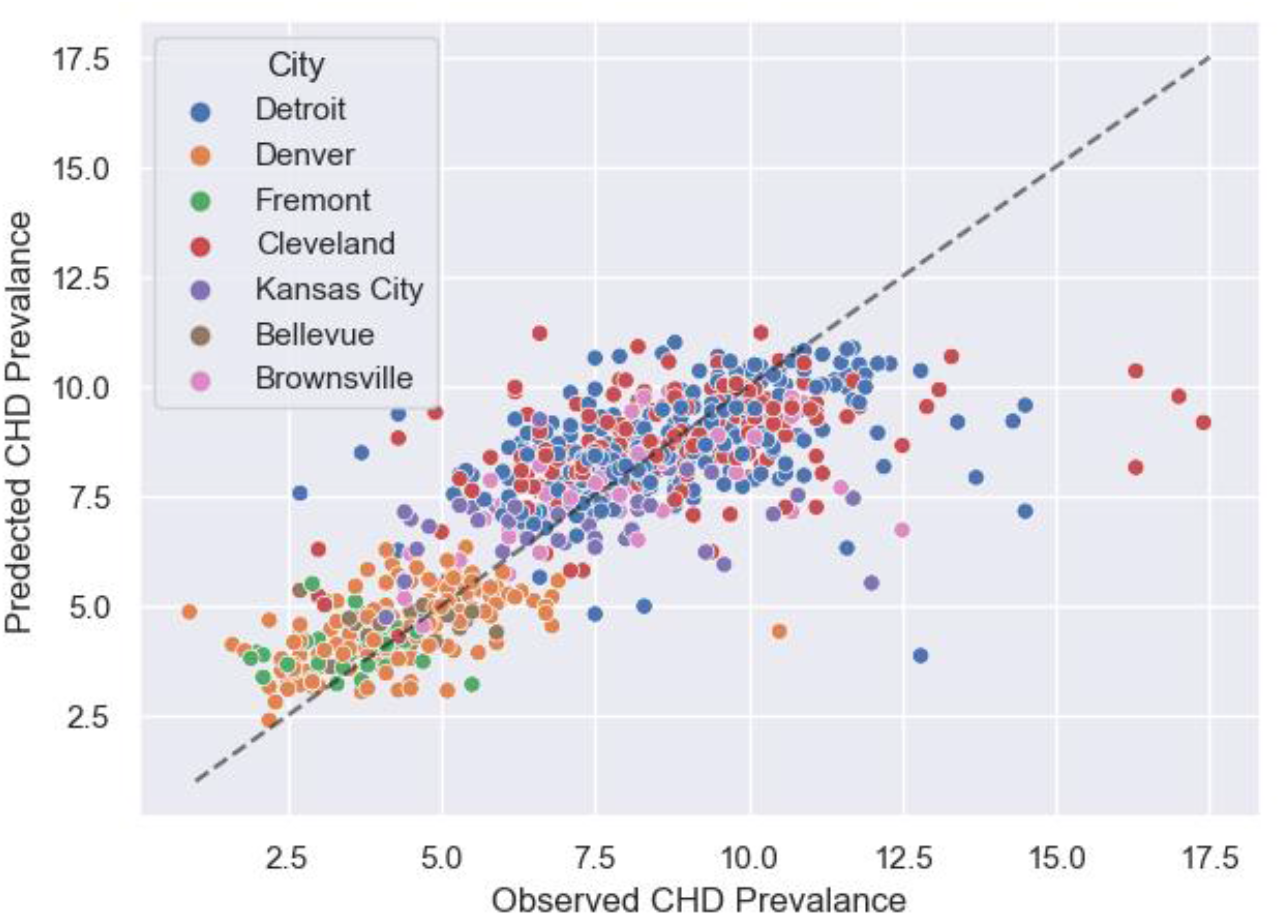
Scatterplot of the actual estimated (observed) and predicted CHD prevalence (in percentage) in seven cities. The Black dotted line represents the *y = x* line.

### Statistical Analysis

#### Features Visualization using Grad-CAM

Elastic net regression models^16^ were used to estimate the census tract-level CHD prevalence by using the DCNN-extracted features from GSV images. There are 4096 features and elastic net can handle this high dimensional data by applying L1 and L2 regularization. Ten-fold cross-validation was repeated 3 times to find the best parameters of the elastic net. Elastic net can select important features by simultaneously performing feature selection and feature shrinkage, so we used it to select top features according to the coefficients of each feature. The top features can be evaluated by examining the magnitudes and signs of their coefficients in the elastic net mode, thus understanding how each feature is associated with CHD prevalence. The top features were then visualized as the saliency map in the original GSV images using Grad-CAM technique^17^, which provides certain explanations of what environmental features the CNN thinks to be associated with neighborhood CHD prevalence.

#### Machine Learning Models with CNN-extracted Features

A variety of machine-learning predictive models were used and compared to explore the association between the CNN-extracted features of GSV images and the tract-level CHD prevalence. The models for this analysis included ExtraTrees regressor (ET), AdaBoost regressor (AB), Random Forest Regressor (RF), Gradient Boosting Regressor (GB), Extreme Gradient Boosting Regressor (XGB), and Light Gradient Boosted Machine Regressor (LGBM). All models were estimated using a 10-fold cross-validation technique for a more robust result. For 10-fold cross-validation, the dataset is split into 10 equal-sized subsets, and the model is trained on 9 subsets and tested on the remaining 1 subset. This process is repeated 10 times until all 10 subsets were used once as the testing set. R-squared values were reported as the measure of association between the CNN-extracted features of GSV images and the tract-level CHD prevalence. The performance of each model was also evaluated using the mean absolute error (MAE) and root mean squared error (RMSE).

#### Multilevel Modeling with Demographics and Socio-Economic Factors

We analyzed the effects of common demographic and socio-economic factors (DSE) as well as CNN-extracted features of GSV images (GSV) associated with the CHD. We built multilevel regression models to account for the effect of these factors including city, age, sex, race, income and education. A multivariate Sparse Partial Least Squares (SPLS) regression^18^ was applied first to the CNN-extracted features to reduce the dimensionality issue and the effect of noise. The selected SPLS components and the demographic and socio-economic factors were then used to fit a Linear Mixed-Effect regression model. Three models were compared in this analysis: 1) a model containing both DSE factors and SPLS components (Combined Model); 2) a model with DSE factors alone (DSE Model); 3) a model with the SPLS components alone (GSV Model). Model performance was assessed using goodness of fits measures such as Likelihood Ratios Tests (as it applied), AIC and BIC criteria. In addition, all models were compared by using R squared values obtained from a Light Gradient Boosted Machine (LGBM) and Random Forest (RF) predictive model, which represent the amount of variance explained by each set of independent variables. The comparison was done using a 10-fold cross validation scheme.

## Results

### Regression results with CNN Features

The 4096 CNN-extracted features from GSV images were able to explain more than 63% of the variance (*R*^2^ = 0.634) on the tract-level CHD prevalence in 7 cities (Figure 1). The ET achieved the best result among all models with the lowest average MAE of 1.11 and RMSE of 1.58. The actual estimate from CDC’s CHD prevalence and the model-predicted CHD prevalence were mapped for all census tracts in 7 cities (Figure 2). There was a good agreement between the actual estimates and predicted CHD prevalence across all census tracts in 7 cities. We found a small number of extreme values that were underestimated by the models in certain census tracts of Detroit and Cleveland. The CHD prevalence of these underestimated census tracts was often more than 12%. When examining the CNN-extracted features using t-SNE, we noticed clustering of census tracts with similar values of CHD prevalence (eFigure2 in the supplement)

**Figure 2.**
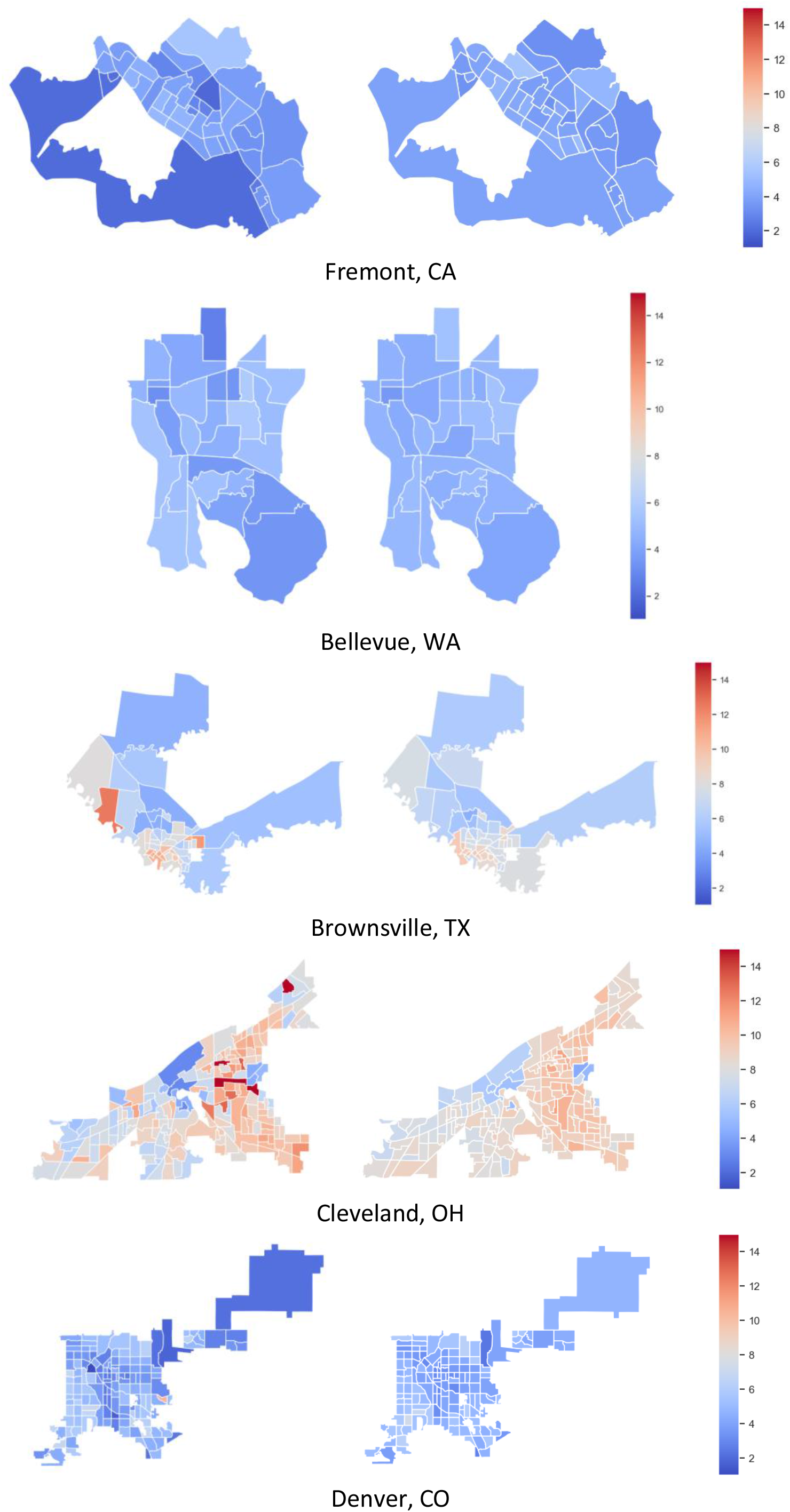
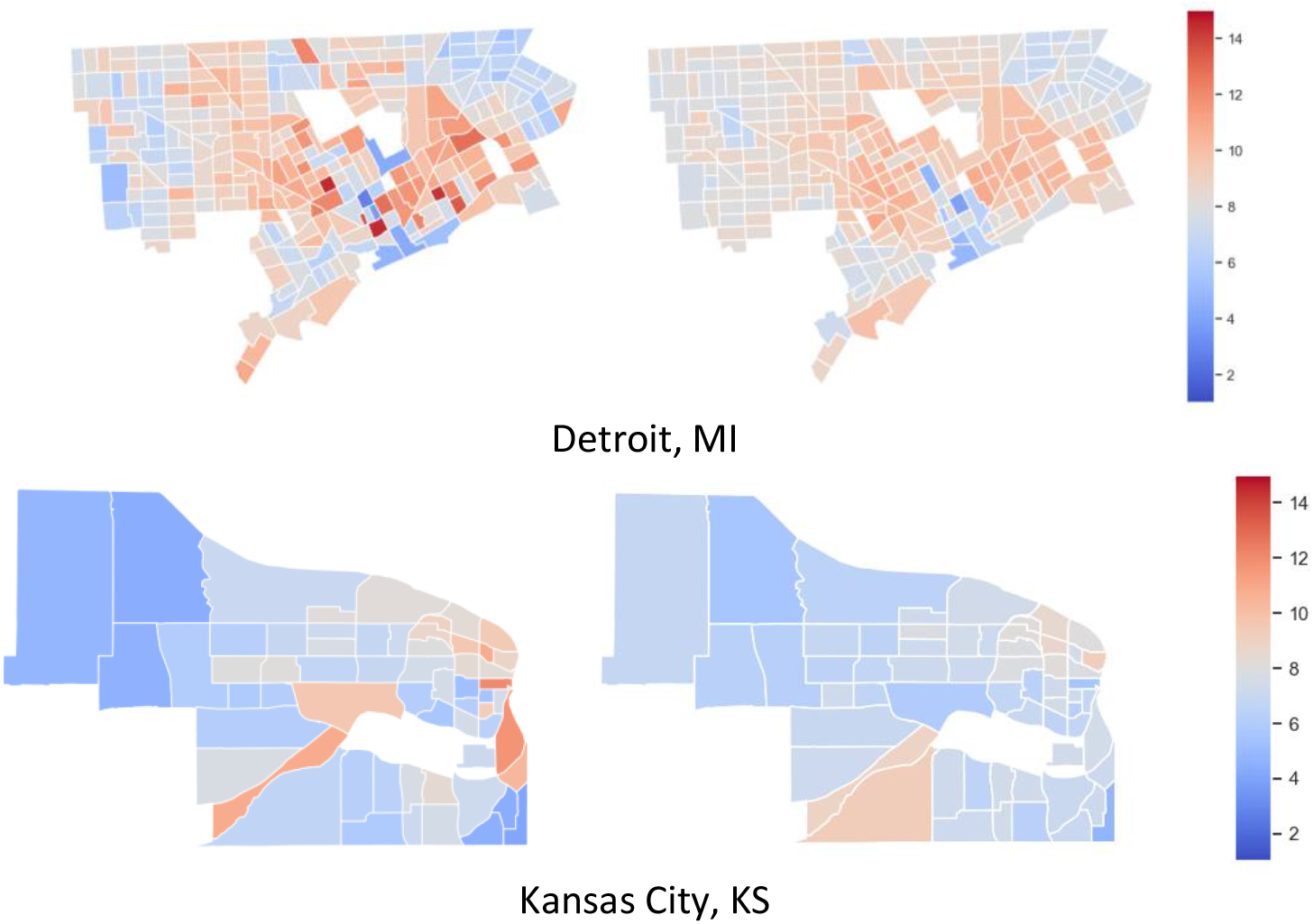
Maps of the actual estimates of cardiovascular heart disease (CHD) prevalence (left) and predicted CHD prevalence (right, in percentage). The predicted CHD prevalence is obtained by averaging the results from 100 random trials based on *k*-fold cross-validation (with *k* = 10).

### Visualization of Top CNN Features

Grad-CAM was utilized to visualize top CNN-extracted features identified from the elastic net model. The saliency maps generated by the Grad-CAM, suggested that feature #1555, which seemed to highlight deteriorated buildings (suggesting neighborhood blight), had a positive association with CHD prevalence (Figure 3a). Another feature (feature #484) that was positively associated with CHD was found to be highlighting road cracks as shown in Figure 3b. In contrast, feature #204 in Figure 3c had a negative association with CHD prevalence, and its heatmap highlighted trees along the road. Feature #1732, seeming to focus on well built houses, also had a negative association with CHD prevalence (Figure 3d).

**Figure 3.**
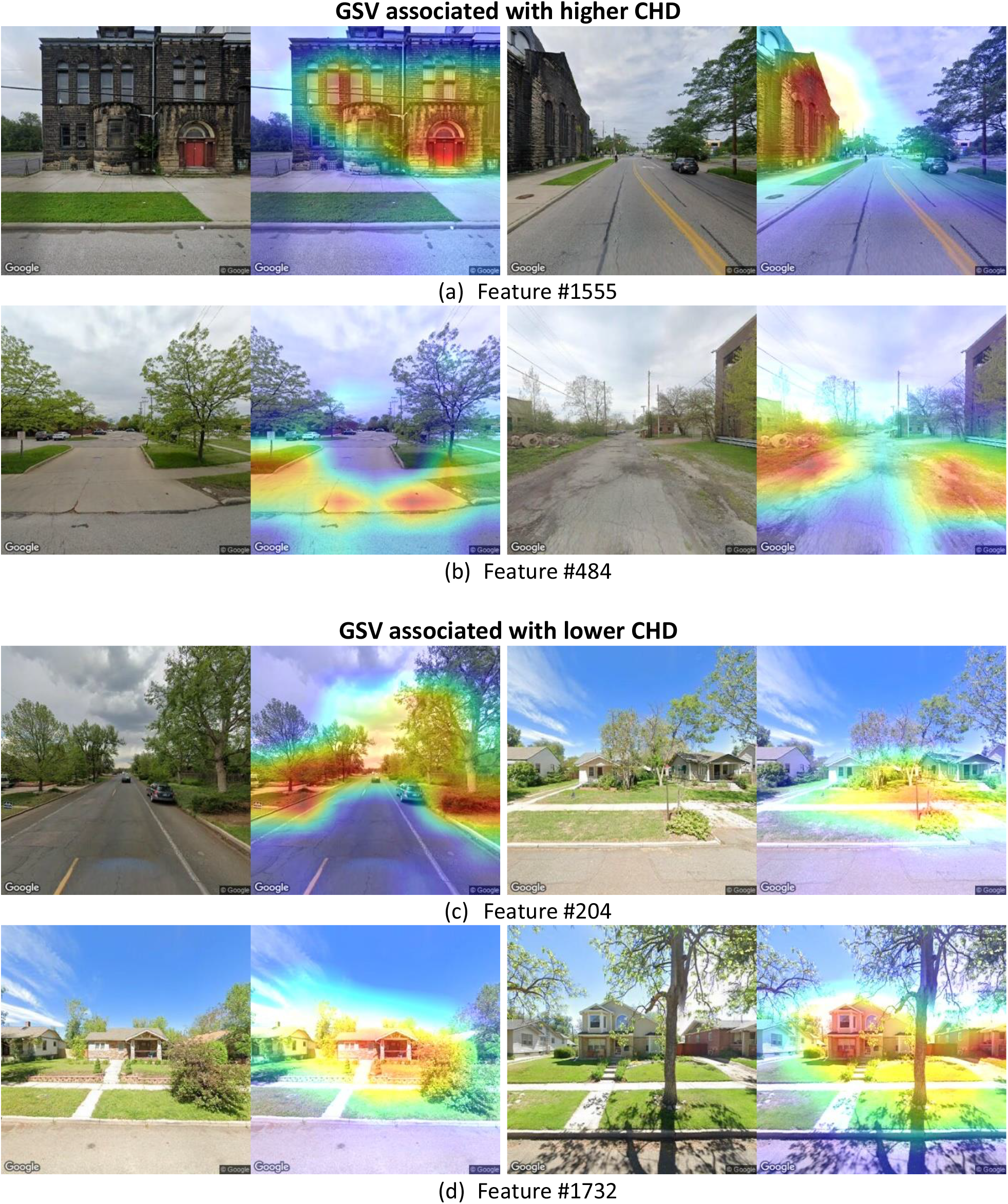
Feature interpretations using Grad-CAMs. Images (a) and (b) show the two pairs of GSV images (left) and their activation maps (right) for the features associated with higher CHD prevalence. Images (c) and (d) show the two pairs of GSV images (left) and their activation maps (right) for the features associated with lower CHD prevalence.

### Comparison of CNN Features with Demographics and Socio-Economic Factors

With SPLS, an optimal model was obtained with *h* = 7 SPLS components (*η* = 0.6), yielding a model with 816 CNN-extracted features that explain 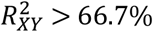 variance of CHD prevalence in the census tracts. All three models were compared with Table 1 shows model comparisons for all three models Likelihood Ratios Tests (LRT, also see eMethods in the Supplement). eTable 1 in the Supplement shows the corresponding regression estimates and ANOVA results. Table 2 and Supplemental eTable 2 show the amount of total explained variance of the GSV models and demographic and socioeconomic (DSE) variables (Table 2: LGBM, eTable 2: RF). After adjusting for each individual variable, we found that the combined model (GSV + DSE) demonstrated a better Goodness of Fit, with statistically significant higher log-likelihood and lower AIC/BIC when compared to GSV or DSE model alone (Table 1). Also, we found that nearly all the SPLS components are statistically significant (eTable 1 in the Supplement). Although the DSE model has lower AIC and BIC values, with a significant LRT, when compared to the GSV model alone (Table 1), the GSV features alone explain more variance of CHD prevalence than the DSE variables (Table 2). Altogether, this indicates that GSV features and traditional demographics and socio-economics variables are both significantly associated and predictive of CHD prevalence.

**Table 1.** AIC, BIC Criteria and Likelihood Ratios Tests (LRT) of LMM models by Model Combination for CHD Prevalence. Models: GSV = the reduced LMM model with only the selected SPLS components CHD: *h* = 7 obtained from the full CNN features; DSE = the reduced LMM model with only the Demographics and Socio-Economic variables; Combined = the full LMM model with both sets of independent variables from GSV and DSE.

| CHD |  |  |  |  |  |  |
| --- | --- | --- | --- | --- | --- | --- |
| LMM Model | AIC | BIC | Log. Lik. | Test | LRT | p-value |
| Combined | 757.0 | 835.0 | -368.0 |  |  |  |
| DSE | 930.0 | 967.0 | -457.0 | Combined vs. DSE | 178.0 | < 1.00E-4 |
| Combined | 757.0 | 835.0 | -368.0 |  |  |  |
| GSV | 984.0 | 1029.0 | -482.0 | Combined vs. GSV | 227.0 | < 1.00E-4 |
| DSE | 930.0 | 967.0 | -457.0 |  |  |  |
| GSV | 984.0 | 1029.0 | -482.0 | DSE vs. GSV | 49.0 | < 1.00E-4 |

**Table 2.** Total Explained Variance of the LGBM Prediction Model by Model Combination for CHD Prevalence.

| CHD | R<sup>2</sup> |  |  |
| --- | --- | --- | --- |
|  | Median | Mean | Std. Error |
| LGBM Model |  |  |  |
| DSE | 0.360 | 0.149 | 0.553 |
| GSV | 0.591 | 0.597 | 0.057 |

## Discussion

While many epidemiological studies have examined associations between cardiovascular disease and individual built environmental features (e.g. greenspace, urban architecture, street connectivity, food availability), our approach focused on machine vision derived physical environment, relying on convolutional neural networks (CNN) and its related techniques to extract features.

Our results showed a good association (*R*^2^ = 0.634) between the CNN-extracted features from GSV and CHD at the census tract level in 7 cities. This indicated that the CNN-extracted features could capture neighborhood features impacting cardiovascular health. The predicted CHD prevalence using CNN-extracted features tended to underestimate in certain areas compared to observed CHD prevalence especially in Detroit and Cleveland. This may suggest that certain CHD-related factors may either not be embedded in these environments at these locations or that perhaps features not captured by street view images, such as demographic factors, ambient factors and other demographic and traditional variables may play a much larger role in these environments.

Our approach took the advantage of the knowledge that fully connected layers in the CNN contain condensed information of the input imagery that can be extracted and utilized for a variety of purposes. We utilized a pre-trained deep convolutional neural network (DCNN) Place365 CNN^15^, so that the deep features from the CNN may be more representative of the built environment. One advantage of this approach is that predefined relevant features in the built environment is not required. The 4096-dimensional deep features embeds all essential information of the built environment in the imagery so that we could retain relevant features as much as reasonably possible. Conversely, the disadvantage of using deep features from a pre-trained CNN is that it becomes difficult to identify corresponding physical features that impact CHD at the neighborhood level. To alleviate this issue and provide certain interpretations of the deep features, we utilized Grad-CAM techniques to visualize the CHD-related features with a saliency map.

Grad-CAM highlighted several potential built environment features that are either associated with higher or lower CHD at the neighborhood level. Deteriorated houses and roads are a feature of urban blight associated with higher CHD. This feature may in turn embody other features in the neighborhood that drive cardiovascular risk, including lack of space for physical activity^7,19^, limited access to nutritionally-balanced food^20^, lack of access to health care^21^. Street greenery on the other hand was highlighted as associated with lower CHD prevalence. This agrees with previous studies that showed a robust association between green space and decreased cardiovascular risks^22,23^.

The results of multilevel modeling using demographics and socio-economic factors, indicate that demographics and socio-economic variables, were still better predictors of CHD prevalence, than GSV features. One explanation is obviously the fact that physical environmental feature even if they represent a “meta” framework for other mediators, may not be sufficient to convey the risk conveyed by other factors which may be sparsely represented. Another reason may be that GSV features may engender increased model complexity, by virtue of including 4096 features (Table 1). However, GSV features alone could still explain preponderant proportion of variance in CHD prevalence (Table 2 and eTable1 in the Supplement). Therefore, by incorporating GSV features into regular DSE variables, one could help improve the prediction of CHD prevalence at the neighborhood level. Our results further suggest that GSV features indeed may be helpful in highlighting specific built environment information related to CHD prevalence at the neighborhood level as illustrated by Grad-CAM methods, which provided a way of identifying built environment information.

There are multiple limitations of this study that should be noted. Firstly, the GSV images used in the study are only available along major streets and roads, and there are some populations who do not live in such neighborhood. However, given the fact that most population live around the urban neighborhood where GSV are abundant, we believe this would not significantly affect the results for majority of census tracts. Further, although Place365 database contains 400+ unique scene categories, it may not include all features that can be found in the built environment. Small objects such as trash, other environmental pollutants and physical domains that may translate into better urban quality of life, may be difficult for computer vision techniques like CNN to detect in a GSV image ^24^. Additionally, the census tracts with CHD prevalence data are from 7 representative U.S cities of CDC PLACES dataset, and may not generalize to all census tracts in the U.S., especially rural areas^25^. Future work is needed to examinate the disparities of urban and rural areas and its cardiovascular-related built environment features.

## Conclusion

Built environment impacts cardiovascular health outcome. In this study, we used Google Street View (GSV) and a scene-pretrained convolutional neural network (CNN) to assess the built environment. We found CNN-extracted features explain significant portion of coronary heart disease (CHD) prevalence at the census tract level. Compared to traditional demographic and socio-economic factors, GSV provides unique information that may relate to CHD such as buildings, greenspace and roads as suggested by the activation maps from Grad-CAM technique. The outcomes of our study provides proof of concept for machine-vision enabled identification of urban network features associated with risk that in principle, may enable rapid identification and targeting interventions in at-risk neighborhoods to reduce cardiovascular burden.

## Data Availability

We confirm that all data referred to in this manuscript are available upon request. Please contact the corresponding author for access to the data.

## SUPPLEMENTARY MATERIAL

### Supplementary Information Text

#### Multilevel Modeling of Demographics and Socio-Economic Effects

Population Demographics and Socio-Economic status variables are well-known factors associated with the prevalence of disease such as those of interest in this study. To assess the contribution of these effects besides the CNN-extracted features to the crude prevalence of CHD a multilevel-level regression model was built to simultaneously account for the effect of City, as well as variables Age (Median), Sex (Female %), Race (White %), Income (Median $), and Education (< High School %) in addition to the 4096 CNN-extracted features. Further, to reduce the dimensionality of the problem and to reduce the effect of noise on the error rates of our inferences, we followed a two-step modeling strategy. In the first step, a multivariate Sparse Partial Least Squares (SPLS) regression^27^ was applied to the CNN-extracted features to consider a reduced set of selected SPLS components (latent variables/factors), to which dimension reduction was further applied by imposing sparsity on their loadings (CNN-extracted features). This was done by using shrinkage estimates of regression coefficients (loadings of component) with combined L1- and L2-penalized estimation. The two tuning parameters of the multivariate Sparse PLS regression model are: (i) the number 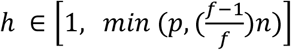 of components that enter the Linear Mixed Effects Model (where *f* is the number of folds and *p* is the dimensionality and *n* is the sample size), and (ii) the sparsity parameter that controls the amount of shrinkage by a combination of the L1- and L2-penalties. Both were tuned simultaneously by 10-fold Cross-Validation. In the second step, a Mixed-Effect regression model was fitted with the first few selected SPLS components augmented with the other Demographics and Socio-Economic variables, all treated here as fixed-effects, and where City was treated as a random effect. Departure from normality of the univariate dependent variable (CHD Prevalence) was tested by EDA analysis and Shapiro-Wilks test. A Box-Cox transformation of the response was applied to minimize departure from normality. Because the dependent variables are continuous, modeling was done by fitting a Linear Mixed Effects Model (LMM). The modeling is entirely supervised because multivariate SPLS regression seeks latent components that not only capture the most variance in the *X*-space (multivariate independent variables) but also the most covariance with the response *Y* (univariate dependent variable of disease prevalence). Finally, AIC and BIC criteria as well as Likelihood Ratios Tests (as it applied) were used for model comparison between the full model and the reduced models with the Demographics and Socio-Economic variables alone or the selected SPLS components alone. Goodness of fit results of all models were also compared by the *R*^2^ amount of explained variance achieved by each set of independent variables in the cross validated Random Forest (RF) and Light Gradient Boosted Machine (LGBM) predictive models as described above.

#### Comparison of CNN Features with Recognized Factors

An optimal model could be obtained after model parameter tuning by cross-validation with a maximum of *h* = 7 SPLS components and *η* = 0.6, yielding a final model comprising *s* = 2031 selected loadings or CNN-extracted features. This resulted in a SPLS model with a cumulative explained variance of CHD: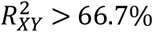. Additional selection of the CNN-extracted features was done by excluding loadings with 0-containing 95% bootstrapped confidence intervals, resulting in a final model of CHD: *s* = 816 loadings. Model specification only included main random and fixed effects and no random interaction effect was found significant. Also, a structure for the covariance matrix was deemed not necessary (not statistically significantly different).

We further refer to the Google Satellite View (GSV) model, as the reduced LMM model where only the selected SPLS components (CHD: *h* = 7) obtained from the full CNN features enter into the model. Likewise, we refer to the Demographics and Socio-Economic (DSE) model, as the reduced LMM model where only the Demographics and Socio-Economic variables enter into the model. We compared the regression estimates and Goodness of Fit (GOF) measures between these reduced models and the Combined LMM model, where both sets of independent variables enter simultaneously. For all three models, Table 1 shows model comparisons by GOF and Likelihood Ratios Tests (LRT), and Supplemental eTable 1 shows the corresponding regression estimates and ANOVA results. Likewise, Table 2 shows the amount of total explained variance by predictive modeling (Table 2: LGBM, Supplemental eTable2: RF).

## Supplementary Figures

**eFigure 1.**
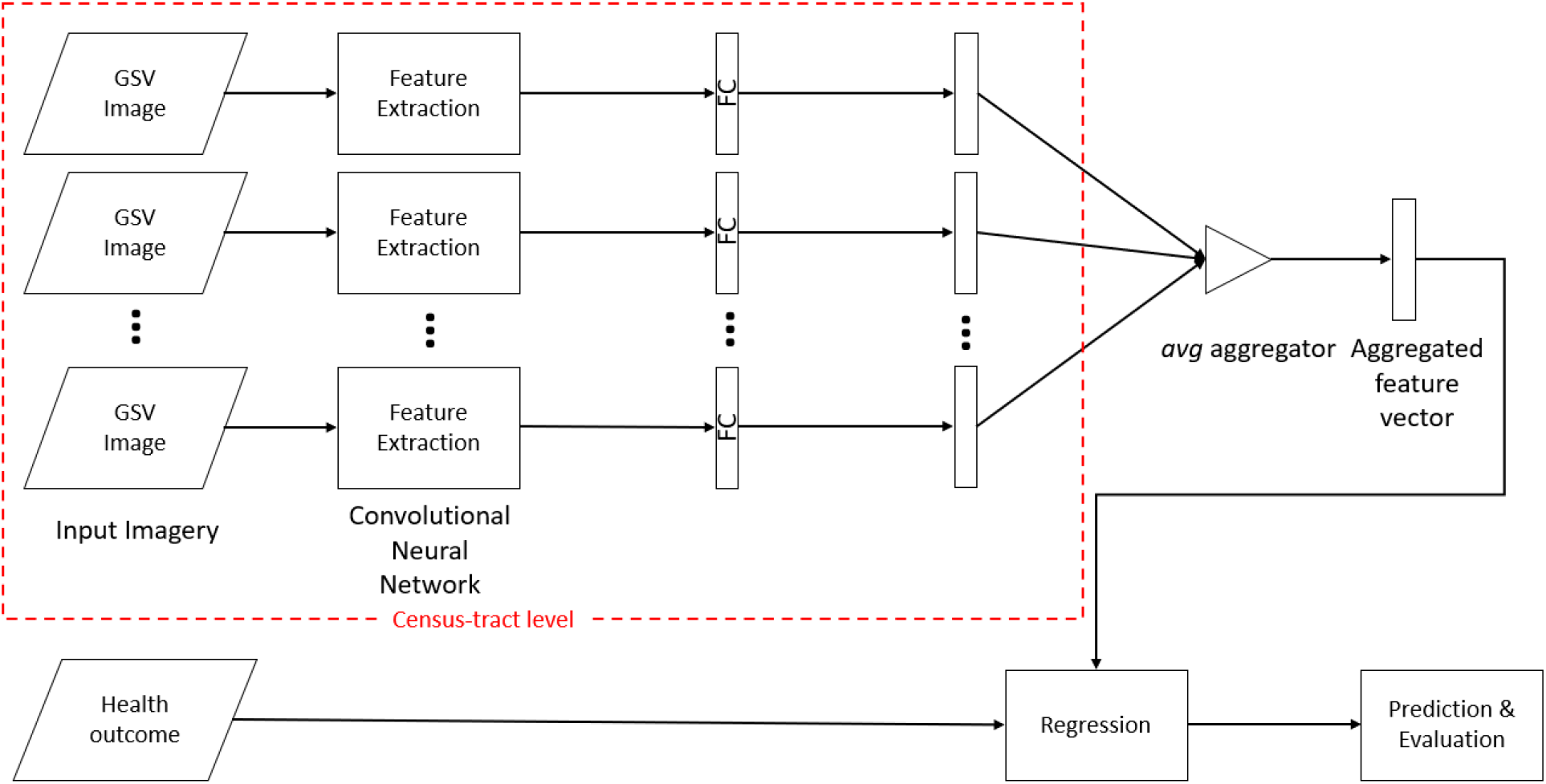
Workflow of the GSV feature extract and regression method. A Places365 pre-trained convolutional neural network – ResNet-50 was used as a feature extractor to obtain the deep features from GSV images. The aggregated feature vector was used in regression to predict CHD prevalence.

**eFigure2.**
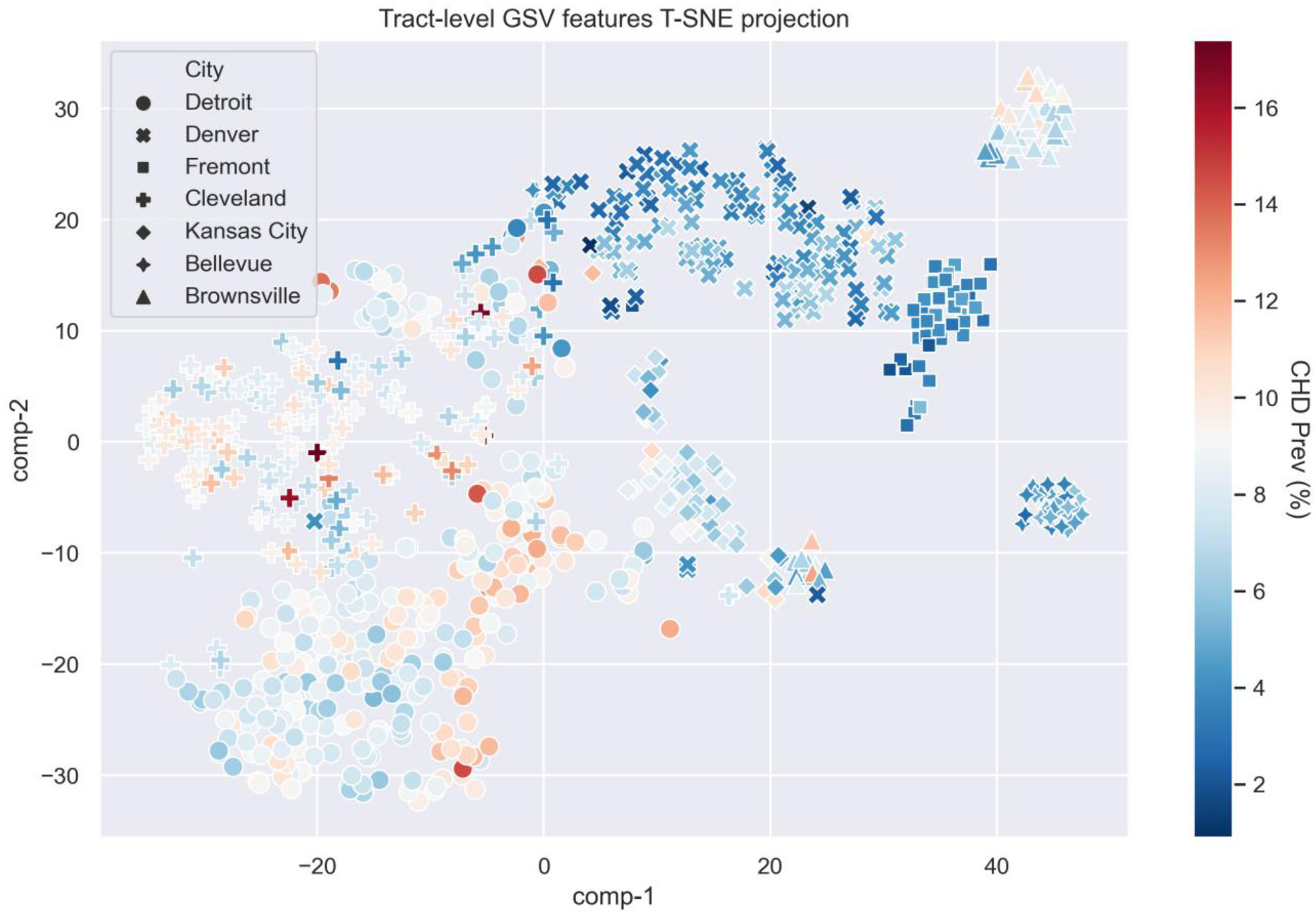
T-SNE projection of the 4096 features from GSV data for the tracts of 7 cities. The projected points are colored by CHD prevalence (%) of the census tract.

## Supplementary Tables

**eTable 1.** Regression Estimates and ANOVA Results by Model Combination for CHD Prevalence.

| CHD |  |  |  |  |  |  |  |
| --- | --- | --- | --- | --- | --- | --- | --- |
|  | Effects | Coeff. | Std. Error | t-value | p-value | F-value | p-value |
| Combined | sex | < 0.1 | < 0.1 | 2.870 | 0.004 | 3428 | < 0.0001 |
|  | Age | < 0.1 | < 0.1 | 14.92 | < 0.001 | 150 | < 0.0001 |
|  | Race | < 0.1 | < 0.1 | -3.450 | 0.001 | 94.000 | < 0.0001 |
|  | Income | < 0.1 | < 0.1 | -10.36 | < 0.001 | 515 | < 0.0001 |
|  | Education | 1.2 | 0.18 | 6.790 | < 0.001 | 132 | < 0.0001 |
|  | Comp.1 | 44.2 | 19.56 | 2.260 | 0.024 | 52.000 | < 0.0001 |
|  | Comp.2 | -23.2 | 11.02 | -2.11 | 0.035 | 6.000 | < 0.0181 |
|  | Comp.3 | 36.5 | 9.65 | 3.780 | < 0.001 | 59.000 | < 0.0001 |
|  | Comp.4 | 32 | 9.95 | 3.220 | 0.001 | 7.000 | 0.0109 |
|  | Comp.5 | -41.2 | 10.75 | -3.83 | < 0.001 | 1.000 | 0.4663 |
|  | Comp.6 | 35.7 | 10.25 | 3.480 | 0.001 | 10 | 0.0014 |
|  | Comp.7 | 49.5 | 10.67 | 4.640 | < 0.001 | 21 | < 0.0001 |
| DSE | Sex | 0.011 | 0.034 | 3.22 | 0.001 | 18 | < 0.0001 |
|  | Age | 0.042 | 0.025 | 17.06 | < 0.001 | 142 | < 0.0001 |
|  | Race | -0.006 | 0.001 | -6.37 | < 0.001 | 85 | < 0.0001 |
|  | Income | 0 | 0 | -10.42 | < 0.001 | 297 | < 0.0001 |
|  | Education | 1.903 | 0.181 | 10.49 | < 0.001 | 110 | < 0.0001 |
| GSV | Comp.1 | 61.8 | 26.19 | 2.36 | 0.019 | 115 | < 0.0001 |
|  | Comp.2 | -25.4 | 13.3 | -1.91 | 0.057 | 54 | < 0.0001 |
|  | Comp.3 | 58.7 | 11.64 | 5.04 | < 0.001 | 137 | < 0.0001 |
|  | Comp.4 | 73.9 | 12.96 | 5.7 | < 0.001 | 23 | < 0.0001 |
|  | Comp.5 | -51.9 | 14.09 | -3.69 | < 0.001 | 5 | 0.0248 |
|  | Comp.6 | 36.2 | 12.74 | 2.86 | 0.004 | 5 | 0.0258 |
|  | Comp.7 | 82.2 | 13.51 | 6.08 | < 0.001 | 37 | < 0.0001 |

**eTable 2.** Total Explained Variance of the RF Prediction Model by Model Combination for CHD Prevalence.

| CHD | R<sup>2</sup> |  |  |
| --- | --- | --- | --- |
|  | Median | Mean | Std. Error |
| RF Model |  |  |  |
| DSE | 0.328 | 0.160 | 0.559 |
| GSV | 0.613 | 0.615 | 0.045 |

